# The Impact of State-Level Prenatal Substance Use Policies on Rates of Maternal and Infant Mortality in the United States: A Legal Epidemiology Study

**DOI:** 10.1101/2022.11.16.22282429

**Authors:** Kathryn A. Thomas, Cara A. Struble, Madeline R. Stenersen, Kelly Moore

**Author notes:** Corresponding Author: Kathryn A. Thomas, The Justice Collaboratory, Yale Law School, SEICHE Center for Health and Justice, Yale School of Medicine.

## Abstract

Rates of maternal and infant mortality remain significantly higher in the United States than other similarly developed countries. Prior research has shown that substance use during pregnancy leads to increased rates of maternal and infant mortality, yet no studies have examined the impact of state-level policies regarding substance use during pregnancy on rates of maternal and infant mortality across all 50 states. The current study utilized publicly available data to examine state-level impact of punitive prenatal substance use laws on maternal and infant mortality. Results revealed that mandated testing laws significantly predicted maternal mortality after controlling for race, poverty, neonatal abstinence syndrome, prenatal care, and substance use in pregnant women, with the entire model accounting for 59.6% of the variance in maternal mortality. Child abuse laws significantly predicted rates of infant mortality, when controlling for race, health insurance, neonatal abstinence syndrome, prenatal care, and substance use in pregnant women, with the entire model accounting for 70.8% of variance in infant mortality. In both regression models, lack of prenatal care increased rates of maternal and infant mortality. Results provide support for the position that laws punishing prenatal substance use may lead to higher rates of maternal and infant mortality. Clinical and policy implications are discussed.

---

Even with pregnancy-related deaths declining worldwide, maternal mortality rates in the United States have continued to rise for the past three decades. In fact, the rates of maternal mortality in the United States more than tripled, rising from 7.2 deaths per 100,000 births in 1987 to 23.8 deaths per 100,000 births in 2020.^1^ An estimated 60% of maternal deaths in the United States are preventable.^2^ Correspondingly, rates of infant mortality remain significantly higher in the United States than other similarly developed countries.^3^

Explaining maternal and infant mortality increases is complicated and likely involves many contributing factors. Research has identified substance use during pregnancy as a major risk factor for both maternal and infant mortality. Prenatal substance use has been shown to be a significant risk factor for both maternal and infant mortality.^4, 5, 6, 7, 8^ From 2007 to 2017, maternal mortality resulting from overdose more than doubled in the U.S.^9, 10^

Although there is a significant relationship between substance use during pregnancy and maternal mortality, researchers have suggested that substance use is also a surrogate for other factors that influence mortality, including reduced prenatal and pediatric healthcare utilization, poor nutrition, unstable housing, and exposure to violence.^11^ It has recently been posited that laws that punish substance use during pregnancy may also lead to increased rates of maternal and infant mortality.^12^ There is an increasing trend in which states have enacted punitive policies regarding substance use during pregnancy with the goal of reducing prenatal substance use, such that the number of states that consider prenatal substance use to be a form of child abuse rose from 12 states in 2000 to 24 states in 2020.^13, 14^ Thirty percent of laws which restrict sexual and reproductive health, including laws punishing prenatal substance use, enacted since 1973 have been established since 2010. ^15^ Research has revealed that fear of seeking care due to social stigma and legal ramifications is one of the most common barriers to prenatal care for pregnant woman with substance use disorders.^16^ To date, there has only been one known study which has examined the impact of prenatal substance use policies on complications in birth.^17^ When reviewing birth records in eight states, Faherty and colleagues found that enactment of punitive policies was associated with significantly greater rates of neonatal abstinence syndrome (NAS), which is a group of conditions caused when an infant withdraws from certain drugs after prenatal exposure.

Although it has been posited that laws punishing substance use during pregnancy could contribute to high rates of maternal and infant mortality, there are no known studies that have examined the impact of state-level policies regarding substance use during pregnancy on maternal and infant mortality across all 50 states. This study utilizes a legal epidemiological approach, which is an emerging area of literature that provides a means to study the effects of laws on health.^18^ This study examined the impact of state-level policies on rates of maternal and infant mortality, including: whether substance use during pregnancy is considered child abuse; whether suspected substance use during pregnancy requires mandated reporting; whether substance use during pregnancy is grounds for civil commitment; and whether substance use during pregnancy requires mandated testing, on rates maternal mortality and infant mortality.

Currently, there are 30 states that have laws defining maternal substance as child abuse, eight states with laws that require mandated testing when maternal substance use is suspected, 25 states with laws that require mandated reporting when maternal substance use is suspected, and three states with laws allowing for civil commitment of pregnant women who use substances. We hypothesized that states with child abuse laws, mandated testing laws, mandated reporting laws, and civil commitment laws will have higher rates of maternal and infant mortality.

Prior work has identified factors associated with maternal and infant mortality identified in prior research, including lack of access to prenatal care, racial disparities, poverty, and lack of access to health insurance. Research has revealed that women who receive no prenatal care are three to four times more likely to experience a pregnancy-related death.^19^ Similarly, research has shown that inadequate prenatal care significantly increases risk of infant mortality, as well as risk of premature birth, still birth, and neonatal death.^20^ Regarding racial disparities, U.S. women who identify as Black, American Indian, and Alaska Native are two to three times more likely to die from pregnancy-related causes than White women. Black, American Indian, and Alaska Native infants are three to five times more likely to die than their Chinese American counterparts.^21^ Laws punishing prenatal substance use are often disproportionately applied, such that low income, Black women who live in Southern states are more likely to be reported by hospital staff, subject to drug testing, charged with a felony, and arrested.^22^ Lack of health insurance and lack of access to healthcare have also been posited to contribute to the high rates of maternal mortality in the United States. Women with Medicaid or no insurance are more likely to receive inadequate prenatal care than women with private insurance, which is known to contribute to maternal mortality. ^23^ Specifically, 84.1 percent of women with private insurance received adequate prenatal care between 2012 and 2014, compared to 64.2 percent of women with Medicaid and 35.7 percent of women with no insurance. ^24, 25^ A body of research in the United States and worldwide has revealed that poverty is a significant predictor of maternal mortality, in part due to differences in rates of prenatal care.^26^

In the current study, we utilize a legal epidemiological approach, hypothesizing that states with child abuse laws, mandated testing laws, mandated reporting laws, and civil commitment laws will have higher rates of maternal and infant mortality. Importantly, we control for a variety of correlates that have been shown in prior research to increase maternal and infant mortality, including race, poverty, and rates of neonatal abstinence syndrome, prenatal care, substance use in pregnant women, health insurance, and prenatal care.

## Methods

### Procedure

Publicly available state-level data were combined from several data sources including: the Guttmacher Institute^1^ state-level coding of whether or not states require mandated testing when pregnant women are suspected of using substances in 2018 (see Tables 1 and 2);^27^ United Health Foundation’ s data on rates of maternal mortality in each state in 2018;^28^ the Centers for Disease Control and Prevention state-level data on infant mortality rates per 100,000 live births in the United States in 2018;^29^ Healthcare Cost and Utilization Project data on state-level rates of neonatal abstinence syndrome (NAS) among newborn hospitalizations in 2018;^30^ Centers for Disease Control and Prevention data on state-level rates of prenatal care in the first trimester in 2018;^31^ U.S. Census state-level estimates of race, poverty levels, and health insurance from 2010 to 2019;^32^ and National Survey on Drug and Alcohol Use (NSDUH) data on substance use during pregnancy in 2018.^33^ Data from all 50 states were included in analyses. All variables were continuous, with the exception of the state-level of coding of prenatal substance use laws, which were coded 1 for the presence of the law and 0 for the lack of the law. See Table 3 for a list of all state-level correlate variables included in the initial analyses.

**Table 1.** State laws on substance use during pregnancy.

| Type of Law | States with that Law |
| --- | --- |
| Child Abuse Laws | AL, AZ, AR, CO, FL, HI, ID, IL, IN, IA, KY, LA, MN, MS, MO, NV, ND, NH, NM, OH, RI, SC, SD, TX, UT, VA, VT, WA, WI, WY |
| Mandated Testing Laws | IN, IA, KY, LA, MN, ND, RI, SD |
| Mandated Reporting Laws | AK, AZ, AR, CA, IL, IA, KY, LA, ME, MA, MI, MN, MT, NV, NH, NJ, ND, OH, OK, PA, RI, SD, VT, VA, WI |
| Civil Commitment Laws | MN, SD, WI |

**Table 2.**
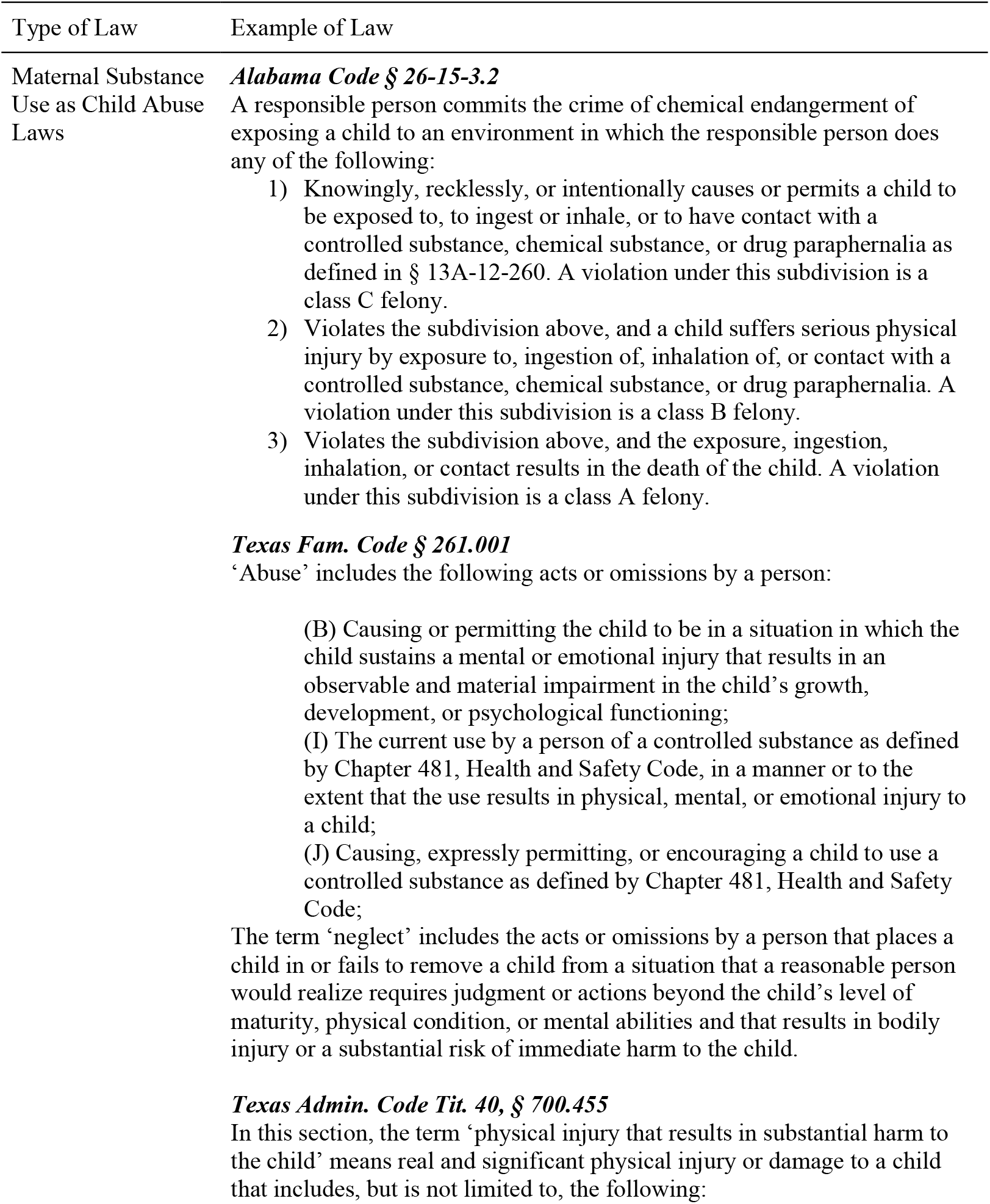
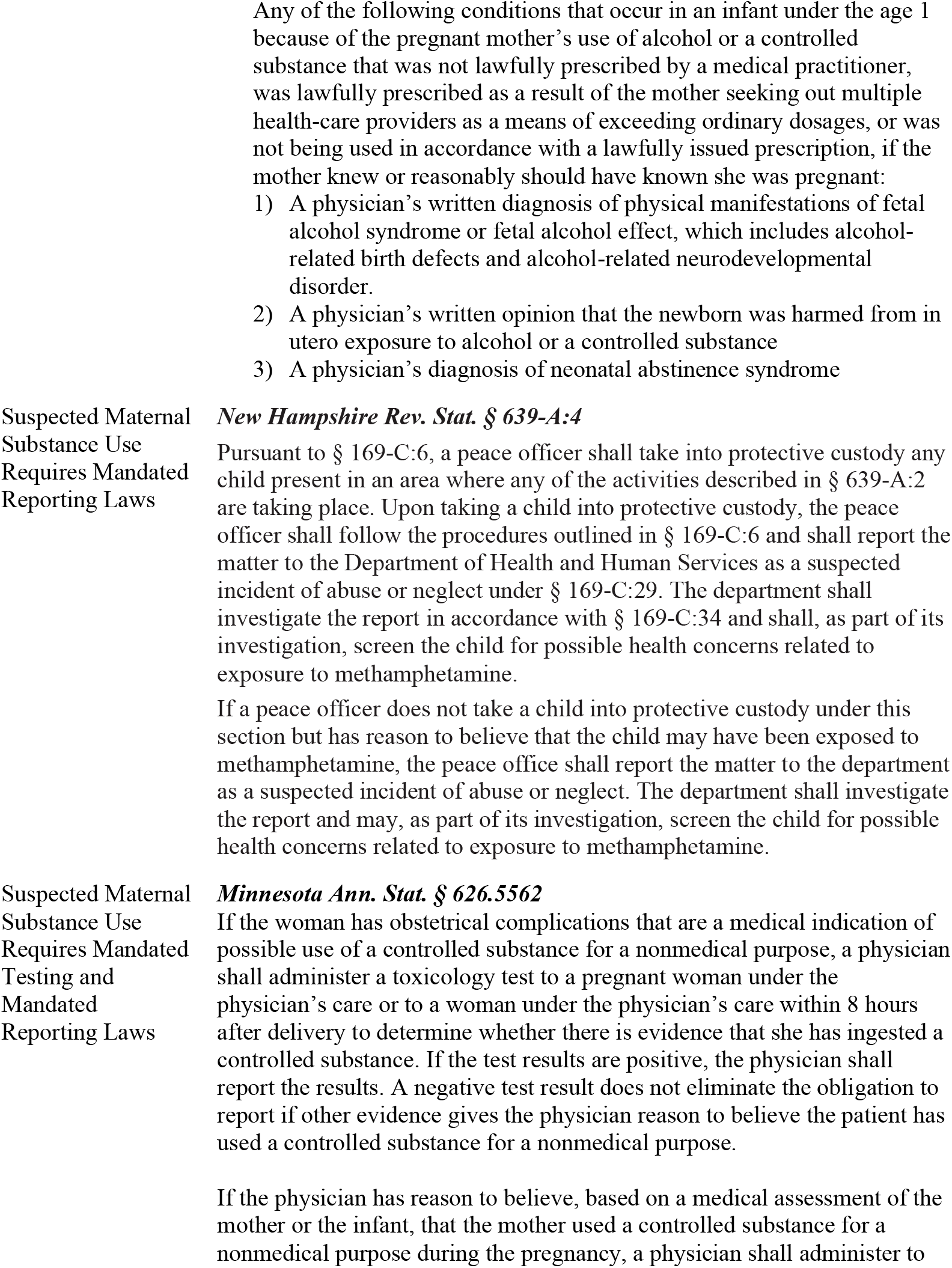
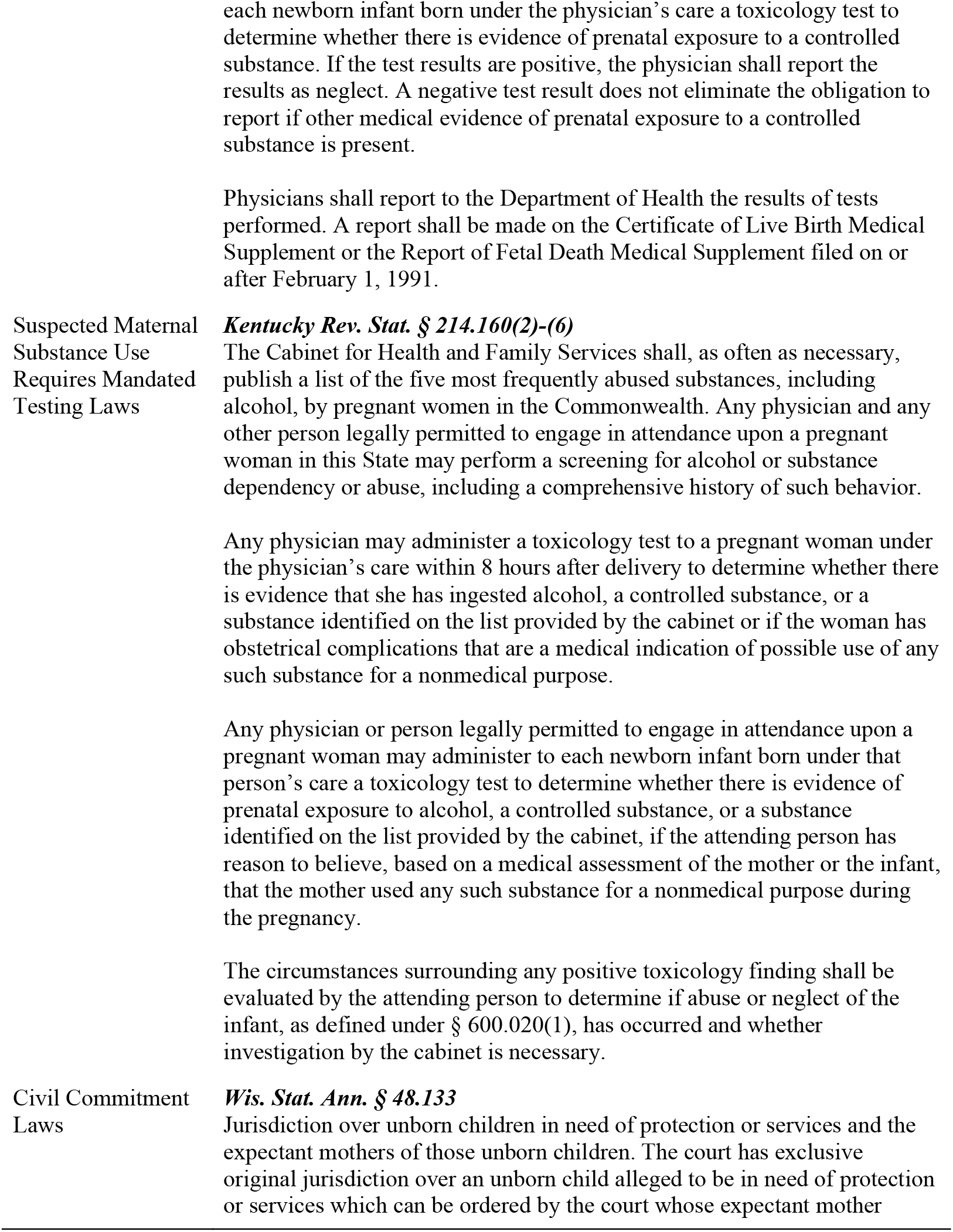
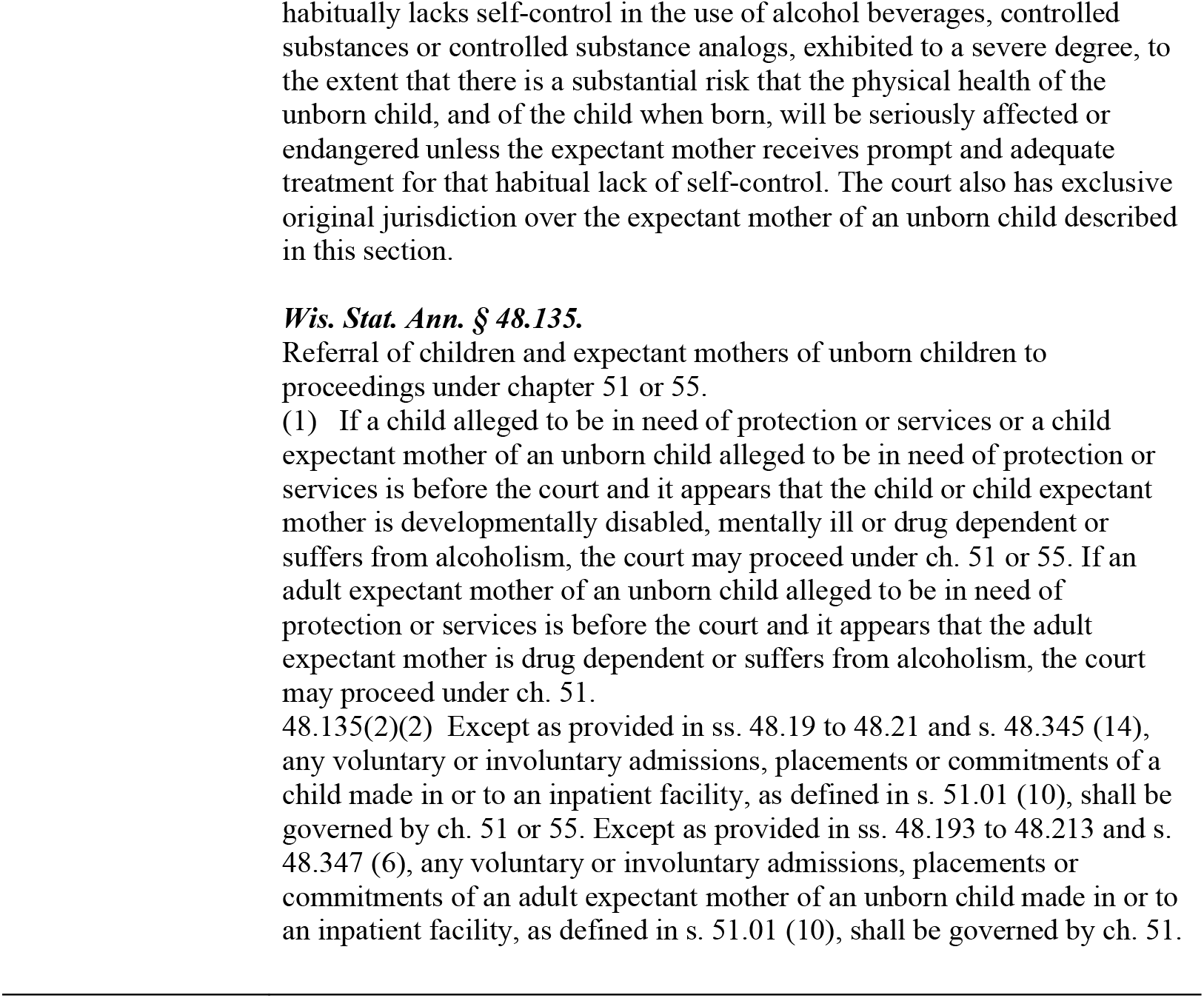
Examples of child abuse laws and mandated testing laws.

**Table 3.** All sources of data included in backward regression models.

| Data Source |
| --- |
| Guttmacher Institute state-level coding of whether states require mandated testing when pregnant women are suspected of using substances in 2018 |
| Center for Disease Control and Prevention state-level data on infant mortality rates per 100,000 live births in the United States in 2018 |
| United Health Foundation's state-level data on maternal mortality rates per 100,000 live births in the United States in 2018 |
| Healthcare Cost and Utilization Project data on state-level rates of Neonatal Abstinence Syndrome (NAS) among newborn hospitalizations in 2018 |
| Center for Disease Control and Prevention data on state-level rates of prenatal care in the first trimester in 2018 |
| U.S. Census state-level estimates of race, poverty levels, percent of state that is rural, and health insurance rates from 2010 to 2019 |
| U.S. Bureau of Labor Statistics state-level rates of unemployment in 2018 |
| National Academy for State Health Policy's coding of whether state's have expanded Medicaid |
| Center for Disease Control and Prevention data on infant mortality in 2018 |
| National Child Abuse and Neglect Data System's data on infants born with exposure to substances in 2018 |
| National Survey on Drug and Alcohol Use (NSDUH) data on substance use during pregnancy in 2018 |

### Data Analysis

Prior to testing the principal hypotheses, bivariate correlations were examined for all variables (see Table 4). A hierarchical multiple regression was performed to examine the state-level impact of laws mandating testing for pregnant women suspected of substance use on maternal mortality. A backward stepwise approach was utilized to select the most salient correlate variables to control for in the first step of the regressions while reducing the likelihood of overfitting the model with variables not contributing to the variance in maternal and infant mortality. The backwards stepwise regression began with a full, saturated model that included all potential demographic variables informed by extant literature. Next, variables that contributed the least amount of variance were removed one-by-one to find a reduced model that best explained the data.

**Table 4.** Correlation Matrix for All Variables.

| Variables | 1 | 2 | 3 | 4 | 5 | 6 | 7 | 8 | 9 | 10 | 11 | 12 | 13 | 14 | 15 |
| --- | --- | --- | --- | --- | --- | --- | --- | --- | --- | --- | --- | --- | --- | --- | --- |
| 1. Maternal Mortality | — |  |  |  |  |  |  |  |  |  |  |  |  |  |  |
| 2. Infant Mortality | .332* | — |  |  |  |  |  |  |  |  |  |  |  |  |  |
| 3. % of Population Identifying as Black | .383** | .533*** | — |  |  |  |  |  |  |  |  |  |  |  |  |
| 4. % of Population with Health Insurance | .009 | .214 | .125 | — |  |  |  |  |  |  |  |  |  |  |  |
| 5. % Below Poverty Line | .333* | .618*** | .436** | -.562*** | — |  |  |  |  |  |  |  |  |  |  |
| 6. % of Neonatal Abstinence Syndrome | -.206 | .097 | -.115 | .468*** | .124 | — |  |  |  |  |  |  |  |  |  |
| 7. % Prenatal Care in First Trimester | -.460*** | .464*** | -.370** | -.111 | -.496*** | .198 | — |  |  |  |  |  |  |  |  |
| 8. % Pregnant Women who used Illicit Drugs, Tobacco, or Alcohol in Past Month | .077 | .272 | .052 | .288* | .135 | .244 | -.054 | — |  |  |  |  |  |  |  |
| 9. % of State that is Rural | .145 | .387** | -.033 | .210 | .263 | .386** | .094 | .079 | — |  |  |  |  |  |  |
| 10. % Infants Born with Substance Exposure | .121 | .182 | .196 | .138 | .261 | .014 | .153 | -.183 | -.018 | — |  |  |  |  |  |
| 11. Unemployment Rates | .039 | .218 | .287* | .515*** | .589*** | .239 | -.323* | .087 | -.131 | .100 | — |  |  |  |  |
| 12. Mandated Testing Laws | .229 | .011 | -.077 | .013 | .006 | -.079 | .092 | .054 | .152 | .339 | -.166 | — |  |  |  |
| 13. Child Abuse Laws | .139 | -.073 | -.073 | -.082 | .093 | -.249 | -.149 | -.171 | .063 | .156 | -.220 | .356* | — |  |  |
| 14. Civil Commitment Laws | -.054 | -.036 | -.152 | -.165 | -.222 | -.148 | .140 | -.065 | .120 | .201 | -.250 | .349* | .206 | — |  |
| 15. Mandated Reporting Laws | -.111 | -.124 | -.231 | .124 | -.137 | -.007 | .245 | .200 | .137 | .291 | .005 | .327* | .082 | .253 | — |

\*\*\* p < .001, \*\*p < .01, \* p < .05, N = 50.

## Results

### Regressions Predicting Maternal Mortality

In terms of zero order correlations, percent of population that identified as Black, percent of population below the poverty line, and percent of women receiving prenatal care in the first trimester were significant predictors of maternal mortality, while the percent of infants with NAS and percent of pregnant women who used illicit drugs, tobacco, or alcohol in the past month were not significant predictors of maternal mortality (see Table 4). The backwards stepwise approach revealed a final regression model which controlled for state-level percent of population that identifies as Black, percent of the population that falls below the poverty line, percent of infants with NAS, percent of women receiving prenatal care in the first trimester, and percent of pregnant women who used illicit drugs, tobacco, or alcohol in the past month. In the first step of the model (which did not include the mandated testing law variable), none of the correlates were significant predictors of maternal mortality. In the second step of the model, the results of the multiple regression revealed that when controlling for correlates identified in the first step, the presence of mandated testing laws was a significant predictor of the rates of maternal mortality. See Table 2 for examples of mandated testing laws. Additionally, the presence of prenatal care in the first trimester became a significant predictor in the second step of the model. See Table 5.

**Table 5.** Regression analyses examining the impact of mandated testing laws on maternal mortality.

| Predictors | *B* | *R*<sup>2</sup> |
| --- | --- | --- |
| *Step 1:* |  |  |
| Percent of Population Identifying as Black | 20.076 | .537** |
| Percent of Women Receiving Prenatal Care First Trimester | -.581 |  |
| Percent Below Poverty Line | .341 |  |
| Percent of Infants with Neonatal Abstinence Syndrome | -.168 |  |
| Percent of Pregnant Women who used Illicit Drugs, Tobacco, or Alcohol in the Past Month | .085 |  |
| *Step 2:* |  |  |
| Percent of Population Identifying as Black | 22.694 | .596** |
| Percent of Women Receiving Prenatal Care First Trimester | -.664* |  |
| Percent Below Poverty Line | .221 |  |
| Percent of Infants with Neonatal Abstinence Syndrome | -.122 |  |
| Percent of Pregnant Women who used Illicit Drugs, Tobacco, or Alcohol in the Past Month | .058 |  |
| Mandated Testing Laws | 6.660* |  |

\*\* p < .01, \* p < .05, N = 50.

Hierarchical multiple regressions including the other three laws affecting substance use in pregnant women (substance use during pregnancy is considered child abuse, suspected substance use during pregnancy requires mandated reporting, and substance use during pregnancy as grounds for civil commitment) revealed that, when controlling for demographic correlates, the three laws were not significant predictors of maternal mortality.

### Regressions Predicting Infant Mortality

In terms of zero order correlations, percent of population that identified as Black and percent of women receiving prenatal care in the first trimester were significant predictors of infant mortality, while the percent of infants with NAS, percent of population with health insurance, and percent of pregnant women who used illicit drugs, tobacco, or alcohol in the past month were not significant predictors of infant mortality (see Table 4). The backwards stepwise approach revealed a final regression which controlled for state-level percent of population that identifies as Black, percent of population receiving prenatal care in the first trimester, percent of the population with health insurance, percent of infants with NAS, and percent of pregnant women who used illicit drugs, tobacco, or alcohol in the past month. In the first step of the model (which did not include the child abuse law variable), only the percent of the population that identifies as Black and the percent of population receiving prenatal care in the first trimester were significant predictors of infant mortality. The results of the multiple regression revealed that, when controlling for correlates identified in the first step, the presence of child abuse laws was a significant predictor of the rates of infant mortality. See Table 2 for examples of child abuse laws. In the second step of the model, percent of population with health insurance became a significant predictor of infant mortality despite not being significant in the first step of the regression. See Table 6.

**Table 6.** Regression analyses examining the impact of child abuse laws on infant mortality.

| Predictors | *B* | *R*<sup>2</sup> |
| --- | --- | --- |
| *Step 1:* |  |  |
| Percent of Population Identifying as Black | 4.740** | .667** |
| Percent of Women Receiving Prenatal Care First Trimester | -.075* |  |
| Percent of Population with Health Insurance | -.002 |  |
| Percent of Infants with Neonatal Abstinence Syndrome | .021 |  |
| Percent of Pregnant Women who used Illicit Drugs, Tobacco, or Alcohol in the Past Month | .026 |  |
| *Step 2:* |  |  |
| Percent of Population Identifying as Black | 5.196** | .708** |
| Percent of Women Receiving Prenatal Care First Trimester | -.079** |  |
| Percent of Population with Health Insurance | -.066* |  |
| Percent of Infants with Neonatal Abstinence Syndrome | .029 |  |
| Percent of Pregnant Women who used Illicit Drugs, Tobacco, or Alcohol in the Past Month | .030 |  |
| Substance Use as Child Abuse Laws | .542* |  |

\*\* $p < .01$ , \* $p < .05$ , $N = 50$ .

Hierarchical multiple regressions including the other three laws impacting substance use in pregnant women (substance use during pregnancy requires mandated testing, suspected substance use during pregnancy requires mandated reporting, and substance use during pregnancy as grounds for civil commitment) revealed that, when controlling for demographic correlates, the three laws were not significant predictors of infant mortality.

## Discussion

This study is the first to examine the impact of state-level policies regarding substance use during pregnancy with maternal and infant mortality across all 50 states. Results newly revealed that the presence of mandated testing laws significantly predicted rates of maternal mortality, when controlling for state-level correlates, including race (Black), poverty, NAS, prenatal care, and substance use in pregnant women, with the entire model accounting for 59.6% of the variance in maternal mortality. Results also newly demonstrated that the presence of child abuse laws significantly predicted rates of infant mortality after controlling for state-level correlates, including race (Black), health insurance, NAS, prenatal care, and substance use in pregnant women, with the entire model accounting for 70.8% of variance in infant mortality. The results of this study add to the findings of Faherty and colleagues, who found that the enactment of policies that punish maternal substance use were associated with significantly greater rates of NAS in eight states. Specifically, this study provides a cross-sectional analysis of the impact of mandating testing laws on maternal mortality and child abuse laws on infant mortality, suggesting that some laws that punish substance use during pregnancy may have an inadvertent impact of increasing maternal and infant mortality.

Researchers have suggested that substance use during pregnancy may increase one’ s chances of receiving inadequate prenatal care, which has been shown to increase both maternal and infant mortality^34, 35^ It is possible that women may avoid prenatal care based on fear of legal consequences. In both hierarchical multiple regression models examining maternal and infant mortality, when controlling for the relevant state-level correlates (including race, health insurance status, poverty rate, rates of NAS, and rates of substance use in pregnant women), lack of prenatal care was a significant predictor of both maternal and infant mortality. This suggests provides preliminary support that reduced rates of prenatal care may be related to greater maternal and infant mortality in light of punitive practices related to substance use. Future research accessing medical records or Medicaid data would be helpful to further explore this potential mechanism.

It is important to note that these findings are cross-sectional and therefore cannot provide support for causal relationships. However, the findings provide preliminary support for the relationship between punitive prenatal substance use laws, reduced rates of prenatal care, and increased rates of maternal and infant mortality. Healthcare providers should help pregnant women who use substances receive prenatal care, rather than punishing their substance use through criminal charges or mandated testing. Policy reform could potentially reduce maternal and infant mortality by reducing the stigma and punishment for pregnant women seeking prenatal care. Further, a multidisciplinary approach, in which prenatal care providers can refer clients to substance use and mental health treatment without fear of repercussions and stigma, has the potential to reduce risk for maternal and infant mortality. Clinicians should engage in collaborative clinical discussions with clients to enhance health equity for both the mother and baby. Clinicians can play an important role in collaborating with their pregnant clients’ obstetricians to ensure that the clients’ physical and mental health care needs are met.

It is also important to note that in the maternal mortality model, race (specifically the percentage of the population that identifies as Black) had a significant zero-order correlation with maternal mortality, such that states with more Black residents had higher rates of maternal mortality (as has been found in prior work.^36, 37, 38, 39^); however, in both the first and second step of the regression model, the percent of the population that identifies as Black was no longer significant, suggesting that there is shared variance between this variable and the other demographic and legal variables in the model (including mandated testing laws, poverty, NAS, prenatal care, and substance use in pregnant women). In our zero-order correlations, states with a higher percentage of people who identify as Black tended to be states with lower levels of prenatal care and higher rates of unemployment and poverty. It is likely that, in these states, Black people are experiencing racial disparities with regards to access to healthcare.^40, 41^ This is consistent with prior research which revealed that Black women are more likely to experience negative pregnancy-related health outcomes and healthcare discrimination, including higher rates of pregnancy-related deaths, lower rates of access to healthcare (including prenatal care), and higher rates of racial discrimination during their prenatal care.^42, 43, 44, 45, 46, 47^ This is also consistent with prior research showing that low income, Black women who live in Southern states are more likely to be targeted and impacted by laws punishing prenatal substance use.^48^

The current study is not without limitations. First, the study was conducted with state-level data from several publicly available federal and state-level datasets. Thus, the results are examining significant trends on the state-level rather than the individual impact of policies on pregnant women’ s experiences with the health care and legal systems. Future research should examine pregnant women’ s individualized experiences with policies affecting pregnant women who use substances, to better understand the stigma or fear of legal repercussions that women may experience. Second, the data utilized in this study were generally collected in the same year, which does not allow for an understanding of how changes in policies may increase or decrease maternal and infant mortality. Further research can utilize a longitudinal approach to explore the impact of changes in policies on rates of maternal and infant mortality. Third, preliminary research has suggested that, within each state, there are inconsistences in how local jurisdictions document substance use during pregnancy and file petitions for court interventions and child removal.^49^ Further research is needed to better understand inconsistencies in how prenatal substance use policies are enforced within each state. Finally, further research is needed to understand the mechanism by which mandated testing laws predict maternal mortality and child abuse laws predict infant mortality, as this study’ s cross-sectional design does not allow for causal inferences.

## Data Availability

All data are available online.

## Footnotes

1 The Guttmacher Institute is a leading nonprofit research and policy institute, founded in 1968, with the goal of advancing sexual and reproductive health and rights throughout the world. The work of the Guttmacher Institute is guided by a 24-member interdisciplinary board of directors, including legal scholars, researchers, and medical professionals.

## References

1 Centers for Disease Control and Prevention. Maternal mortality rates in the United States. https://www.cdc.gov/nchs/data/hestat/maternal-mortality/2020/maternal-mortality-rates-2020.htm#:~:text=In%202020%2C%20861%20women%20were,20.1%20in%202019%20(Table). (2022, accessed 9 April 2022).

2 Petersen EE, Davis NL, Goodman D, et al. Vital signs: Pregnancy-related deaths, United States, 2011–2015, and strategies for Prevention, 13 states, 2013–2017. MMWR Morbidity and Mortality Weekly Report. 2019; 68. DOI: 10.15585/mmwr.mm6818e1.

3 MacDorman MF, Mathews TJ, Mahongoo AD, Zeitlin J. International Comparisons of Infant Mortality and Related Factors: United States and Europe, 2010. National Vital Statistics Reports. 2014. 63:175–182. doi:10.1111/j.1523-536x.2006.00102.x

4 Virginia Department of Health, Virginia Maternal Mortality Review Team. Pregnancy-Associated Deaths From Drug Overdose in Virginia, 1999-2007. https://www.vdh.virginia.gov/content/uploads/sites/18/2016/04/Final-Pregnancy-Associated-Deaths-Due-to-Drug-Overdose.pdf. (2015, accessed 15 September 2020).

5 Texas Department of State Health Services. Joint Biannual Report. https://dshs.texas.gov/mch/pdf/2016BiennialReport.pdf. (2016, accessed 3 February 2021).

6 Maryland Department of Health and Mental Hygiene. Maryland maternal mortality review 2016 annual report. http://healthymaryland.org/wp-content/uploads/2011/05/MMR_Report_2016_clean-copy_FINAL.pdf. (2016, accessed 2 February 2021).

7 Wolfe EL, Davis T, Guydish J, and Delucchi KL. Mortality Risk Associated with Perinatal Drug and Alcohol Use in California. Journal of Perinatology. 2005. 25:93–100. doi:10.1038/sj.jp.7211214

8 Burd L and Wilson H. Fetal, infant, and child mortality in a context of alcohol use. American Journal of Medical Genetics. 2004. 127: 51–58.

9 Cleveland LM, McGlothen-Bell K, Scott LA, et al. A Life-course theory exploration of opioid-related maternal mortality in the United States. Addiction. 2020. 115: 2079–2088.

10 Centers for Disease Control and Prevention. Substance Use During Pregnancy. https://www.cdc.gov/reproductivehealth/maternalinfanthealth/substance-abuse/substance-abuse-during-pregnancy.htm#:~:text=Opioid%20use%20disorder%20during%20pregnancy,neonatal%20abstinence%20syndrome%20(NAS) (2022, accessed 10 April 2022).

11 Wolfe EL, Davis T, Guydish J, and Delucchi KL. Mortality Risk Associated with Perinatal Drug and Alcohol Use in California. Journal of Perinatology. 2005. 25:93–100. doi:10.1038/sj.jp.7211214

12 Faherty LJ, Kranz AM, Russell-Fritch J, Patrick SW, Cantor J, and Stein BD. Association of Punitive and Reporting State Policies Related to Substance Use in Pregnancy With Rates of Neonatal Abstinence Syndrome. JAMA Network Open. 2019. 2:11. doi:10.1001/jamanetworkopen.2019.14078

13 Guttmacher Institute. Substance Use During Pregnancy. https://www.guttmacher.org/state-policy/explore/substance-use-during-pregnancy. (2021, accessed 12 March 2021).

14 Dailard C and Nash E. State Responses to Substance Abuse Among Pregnant Women. Guttmacher Institute. http://www.guttmacher.org/pubs/tgr/03/6/gr030603.html. (2016, accessed 24 July 2020).

15 Hui K, Angelotta C, Fisher CE. Criminalizing substance use in pregnancy: Misplaced priorities. Addiction. 2017;112(7):1123–5.

16 Gopman S. Prenatal and postpartum care of women with substance use disorders. Obstetrics and Gynecology Clinics of North America. 2014;41(2):213–28.

17 ^‘^Faherty LJ, Kranz AM, Russell-Fritch J, Patrick SW, Cantor J, and Stein BD. Association of Punitive and Reporting State Policies Related to Substance Use in Pregnancy With Rates of Neonatal Abstinence Syndrome. JAMA Network Open. 2019. 2:11. doi:10.1001/jamanetworkopen.2019.14078

18 Ramanathan T, Hulkower R, Holbrook J, Penn M. Legal epidemiology: the science of law. J Law Med Ethics. 2017;45(1 suppl):69–72. doi: 10.1177/1073110517703329

19 U.S. Department of Health and Human Services, Office on Women’ s Health. Prenatal Care. https://www.womenshealth.gov/a-z-topics/prenatal-care. (2019, accessed 4 October 2020).

20 Partridge S, Balayla J, Holcroft C, et al. Inadequate prenatal care utilization and risks of infant mortality and poor birth outcome: A retrospective analysis of 28,729,765 U.S. deliveries over 8 years. American Journal of Perinatology. 2021. 29: 787–794.

21 Singh GK and Yu SM. Infant mortality in the United States, 1915-2017: Large social inequalities have persisted for over a century. International Journal of Maternal and Child Health and AIDS (IJMA). 2019; 8: 19–31.

22 Paltrow LM, Flavin J. Arrests of and forced interventions on pregnant women in the United States, 1973–2005: Implications for women’ s legal status and public health. Journal of Health Politics, Policy and Law. 2013;38(2):299–343.

23 Access in Brief: Pregnant Women and Medicaid. Report to Congress on Medicaid and CHIP https://www.macpac.gov/wp-content/uploads/2018/11/Pregnant-Women-and-Medicaid.pdf (2018, accessed 4 October 2020).

24 Access in Brief: Pregnant Women and Medicaid. Report to Congress on Medicaid and CHIP https://www.macpac.gov/wp-content/uploads/2018/11/Pregnant-Women-and-Medicaid.pdf (2018, accessed 4 October 2020).

25 VA Office of Women’ s Health. https://www.womenshealth.gov/a-z-topics/prenatal-care. (2019, accessed 12 March 2021).

26 Ronsmans C, Graham WJ. Maternal mortality: Who, when, where, and why. The Lancet. 2006;368(9542):1189–200.

27 Guttmacher Institute. Substance Use During Pregnancy. https://www.guttmacher.org/state-policy/explore/substance-use-during-pregnancy. (2021, accessed 12 March 2021).

28 United Health Foundation. Health of Women and Children. America’ s Health Rankings https://www.americashealthrankings.org/explore/health-of-women-and-children/measure/overall_hwc_2020/state/ALL (2021, accessed 1 February 2021).

29 Centers for Disease Control and Prevention. Stats of the States - Infant Mortality. https://www.cdc.gov/nchs/pressroom/sosmap/infant_mortality_rates/infant_mortality.htm. (2018, accessed 1 September 2020).

30 Healthcare Cost and Utilization Project. Neonatal abstinence syndrome (NAS) among newborn hospitalizations. NAS Hospitalizations Map - HCUP Fast Stats. https://www.hcup-us.ahrq.gov/faststats/NASMap. (2020, accessed 4 April 2021).

31 Osterman MJK and Martin, JA. Timing and Adequacy of Prenatal Care in the United States, 2016. National Vital Statistics Reports. 2018. 67:1–14.

32 U.S. Census Bureau. Explore Census Data. https://data.census.gov/cedsci/. (2020, accessed 4 October 2020).

33 U.S. Department of Health and Human Services, Substance Abuse and Mental Health Services Administration, Center for Behavioral Health Statistics and Quality. National Survey on Drug Use and Health. 2018. Retrieved from https://datafiles.samhsa.gov/ (2019, accessed 4 October 2020).

34 Wolfe EL, Davis T, Guydish J, and Delucchi KL. Mortality Risk Associated with Perinatal Drug and Alcohol Use in California. Journal of Perinatology. 2005. 25:93–100. doi:10.1038/sj.jp.7211214

35 Maternal Mortality Review Information Application. Enhancing reviews and surveillance to eliminate maternal mortality. Centers for Disease Control and Prevention. https://www.cdc.gov/reproductivehealth/maternal-mortality/erase-mm/mmr-data-brief.html (2019, accessed October 4, 2020).

36 Noursi S, Clayton JA, Bianchi DW, et al. Maternal morbidity and mortality. Journal of Women’ s Health. 2021. 30: 145–146.

37 Singh, GK. Trends and social inequalities in maternal mortality in the United States, 1969-2018. International Journal of Maternal and Child Health and AIDS (IJMA). 2020. 10:29– 42. https://doi.org/10.21106/ijma.444.

38 Tucker MJ, Berg CJ, Callaghan WM, et al. The black–white disparity in pregnancy-related mortality from 5 conditions: Differences in prevalence and case-fatality rates. American Journal of Public Health. 2007; 97: 247–251.

39 Partridge S, Balayla J, Holcroft C, et al. Inadequate prenatal care utilization and risks of infant mortality and poor birth outcome: A retrospective analysis of 28,729,765 U.S. deliveries over 8 years. American Journal of Perinatology. 2021. 29: 787–794.

40 Howell EA, Egorova N, Balbierz A, Zeitlin J, and Hebert PL. Black-white differences in severe maternal morbidity and site of care. American Journal of Obstetrics and Gynecology. 2016. 214:1. doi:10.1016/j.ajog.2015.08.019

41 Partridge S, Balayla J, Holcroft C, et al. Inadequate prenatal care utilization and risks of infant mortality and poor birth outcome: A retrospective analysis of 28,729,765 U.S. deliveries over 8 years. American Journal of Perinatology. 2021. 29: 787–794.

42 Singh, GK. Trends and social inequalities in maternal mortality in the United States, 1969-2018. International Journal of Maternal and Child Health and AIDS (IJMA). 2020. 10:29– 42. https://doi.org/10.21106/ijma.444.

43 Tucker MJ, Berg CJ, Callaghan WM, et al. The black–white disparity in pregnancy-related mortality from 5 conditions: Differences in prevalence and case-fatality rates. American Journal of Public Health. 2007; 97: 247–251.

44 Howell EA, Egorova N, Balbierz A, Zeitlin J, and Hebert PL. Black-white differences in severe maternal morbidity and site of care. American Journal of Obstetrics and Gynecology. 2016. 214:1. doi:10.1016/j.ajog.2015.08.019

45 Partridge S, Balayla J, Holcroft C, et al. Inadequate prenatal care utilization and risks of infant mortality and poor birth outcome: A retrospective analysis of 28,729,765 U.S. deliveries over 8 years. American Journal of Perinatology. 2021. 29: 787–794.

46 Noursi S, Clayton JA, Bianchi DW, et al. Maternal morbidity and mortality. Journal of Women’ s Health. 2021. 30: 145–146.

47 Vintzileos AM, Ananth CV, Smulian JC, Scorza WE, and Knuppel RA. The impact of prenatal care on neonatal deaths in the presence and absence of antenatal high-risk conditions. American Journal of Obstetrics and Gynecology. 2002. 186:1011–1016. doi:10.1067/mob.2002.122446

48 Paltrow LM, Flavin J. Arrests of and forced interventions on pregnant women in the United States, 1973–2005: Implications for women’ s legal status and public health. Journal of Health Politics, Policy and Law. 2013;38(2):299–343.

49 Ondersma SJ, Simpson SM, Brestan EV, and Ward M. Prenatal Drug Exposure and Social Policy: The Search for an Appropriate Response. Child Maltreatment. 2000. 5:93–108. doi:10.1177/1077559500005002002

